# Potentially Modifiable Dementia Risk Factors in Canada: An Analysis of Canadian Longitudinal Study on Aging with a Multi-Country Comparison

**DOI:** 10.1101/2024.02.20.24303090

**Authors:** Surim Son, Mark Speechley, Guangyong Zou, Miia Kivipelto, Francesca Mangialasche, Howard Feldman, Howard Chertkow, Sylvie Belleville, Haakon Nygaard, Vladimir Hachinski, F Pieruccini-Faria, Manuel Montero-Odasso

## Abstract

**BACKGROUND:** Previous estimates suggested that up to 40% of dementia cases worldwide are associated with modifiable risk factors, however, these estimates are not known in Canada. Furthermore, sleep disturbance, an emerging factor, has not been incorporated into the life-course model of dementia prevention.

**Objective:** To estimate the population impact of 12 modifiable risk factors in Canadian adults including sleep disturbance, by sex and age groups, and to compare with other countries.

**Design:** Cross-sectional analysis of Canadian Longitudinal Study on Aging Baseline data

**Setting:** Community

**Participants:** 30,097 adults aged 45 years and older.

**Measurements:** Prevalence and Population Attributable Fraction (PAF) of less education, hearing loss, traumatic brain injury, hypertension, excessive alcohol, obesity, smoking, depression, social isolation, physical inactivity, diabetes, and sleep disturbance.

**Results:** The risk factors with the largest PAF were later life physical inactivity (10.2%; 95% CI, 6.8% to 13%), midlife hearing loss (6.5%; 3.7% to 9.3%), midlife obesity (6.4; 4.1% to 7.7%), and midlife hypertension (6.2%; 2.7% to 9.3%). The PAF of later life sleep disturbance was 3.0% (95% CI, 1.8% to 3.8%). The 12 risk factors accounted for 51.9% (32.2% to 68.0%) of dementia among men and 52.4% (32.5% to 68.7%) among women. Overall, the combined PAF of all risk factors was 49.2% (31.1% to 64.9%), and it increased with age.

**Conclusion:** Nearly up to 50% of dementia cases in Canada could be prevented by modifying the 12 risk factors across the lifespan. Canadian risk reduction strategies should prioritize targeting physical inactivity, hearing loss, obesity, and hypertension.

## INTRODUCTION

With rapid global population aging, the number of individuals living with dementia worldwide is expected to triple, from 57 million to 152 million, by 2050.^1^ In Canada, dementia prevalence is projected to increase by 66% by 2031.^2^ Dementia is a multifactorial syndrome that results from multiple pathologies including those that cause neurodegeneration as well as vascular, metabolic and inflammatory processes that are associated with potentially modifiable risk factors.^3,4^ Lifestyle interventions offer a promising non-pharmacological approach to reducing dementia burden by tempering modifiable risk factors. Risk reduction can potentially be reached through individual and public health approaches, which could complement emerging disease-modifying treatments directed at the pathological processes.^4^

The 2020 Lancet Commission Report on Dementia Prevention, Intervention, and Care^5^ indicated that up to 40% of dementia cases worldwide are attributable to 12 modifiable factors comprising health behaviours, illnesses, and environmental exposures across the lifespan, known as the life course model of dementia prevention. This estimation is based on the weighted population attributable fraction (PAF), which quantifies the contribution of a given risk factor to dementia by combining both prevalence and the association between risk factor and disease, such as risk ratio, while adjusting for intercorrelation among risk factors.

Following the Lancet series,^5,6^ the population impact of dementia risk factors has been estimated in other countries with differences in risk factor profiles.^7-12^ Given that the same analytical approach and risk ratio were used to estimate PAF in all the studies, the differences in PAF across countries are mainly driven by differences in risk factor prevalence. For instance, less education is a greater contributing factor to dementia than social isolation in low-income countries as they tend to have stronger social ties and social support but have limited access to education compared to high-income countries.^9^ Despite being one of the high-income countries with a universal health care system, Canada now more than ever needs dementia risk reduction strategies, as it is reaching the super-aged country status, where it has ≥ 20% of its population composed of older adults.^13^ However, no studies have estimated the population impact of the life-course model of modifiable risk factors for dementia in Canada, which is crucial for guiding evidence-based decision-making in the development of reduction strategies.

Besides the 12 risk factors included in the life course model of dementia prevention, the Lancet Commission Reports also identified sleep and diet as emerging risk factors.^5,6^ Given the number of intervention trials delivering sleep and diet interventions to improve cognition, it is timely to quantify their population impact.^14-16^ Estimating the population impact of diet is challenging due to the difficulty in measuring the diet pattern. For sleep, a recent study estimated that 5.7% of dementia cases are attributable to unhealthy sleep duration, using UK Biobank data.^17^ However, the population impact of a sleep disturbance encompassing other conditions has not been estimated nor included in the life course model of dementia prevention.

Although air pollution is included in the life-course model^5^, focusing only on lifestyle risk factors, rather than including an environmental risk factor, would be more beneficial in guiding the dementia risk reduction program.

Therefore, to provide evidence to help inform future lifestyle interventions in Canada, we aimed to estimate the prevalence and potential population impact of modifiable risk factors, including sleep disturbance, in Canadian adults using the largest Canadian population cohort study, and to compare the contribution of modifiable risk factors with other countries. To further provide evidence for tailoring prevention strategies, we aimed to estimate these measures by age groups and sex. Since the PAF of dementia risk factors has not been estimated in Canada, this study is much needed to tailor and develop Canada’s dementia risk reduction program.

## METHODS

This was a cross-sectional analysis of baseline data from the Canadian Longitudinal Study on Aging (CLSA) Comprehensive cohort. We followed STROBE guidelines for a cross-sectional study and conducted analyses using the *survey* and *psy* package in R Version 4.2.0.

### Data source

CLSA is a prospective cohort study following 51,388 Canadians aged 45-85 years at recruitment for 20 years.^18^ Participants were recruited to either Tracking or Comprehensive cohorts, which differed in sampling methods and data availability, as described elsewhere and in Figure S1 in Appendix.^18^ The comprehensive cohort was used as it included participants who had undergone detailed and comprehensive face-to-face assessments that included a neuro-cognitive battery, sensory-vision, audiometry, proprioception testing as well as physical and mobility assessments. Individuals with a diagnosis of cognitive impairment and or dementia, full-time members of the armed forces, residents of First Nations reserves, territories, or long-term care institutions (only those that provide 24 hours of nurse care), and those who could not respond in English or French at recruitment were excluded.^18^ Baseline data were collected from 2012 to 2015.^18^

### Risk factors

A total of 12 risk factors were identified using operational definitions from the Lancet report^5^ and 2 initial PAF estimation studies for dementia.^19,20^ Less education was defined as having less than secondary school graduation. Hearing loss was derived from an average hearing level of >25 dB at 500, 1000, 2000 and 4000 Hz in the better ear.^21^ Traumatic brain injury was defined as having at least one head injury caused by a vehicular crash, fall, or sports-related activities that resulted in losing consciousness. An average systolic blood pressure ≥140 mmHg across six seated measurements excluding the first reading or self-reported diagnosis was used to define hypertension. The number of drinks per week was converted to the unit of alcohol using the alcohol unit conversion formula by UK National Health Service, 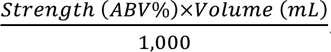.^22^ Based on the converted unit of alcohol, >21 units was used to indicate excessive alcohol use. BMI of ≥30 kg/m^2^ was used to categorize obesity. Daily or occasional cigarette smoking in the past 30 days was used to represent current cigarette smoking. Depression was categorized based on the self-reported diagnosis of clinical depression. Social isolation was defined as having less than one social contact within a month with family, friends, or neighbours. The level of physical activity was measured using the Physical Activity Scale for the Elderly (PASE) questionnaire. The total hours of physical activity per week were computed using the lowest point of each frequency and duration category with the exception that the mid-point was used for the lowest frequency category. Based on the estimated minutes of moderate-to-vigorous physical activity, physical inactivity was defined as <150 minutes. A self-report diagnosis of type 2 diabetes by a physician was used for diabetes. We characterized sleep disturbance based on the definition used in the systematic review^23^ from which we used their reported relative risk for PAF calculation. Sleep disturbance was broadly defined to encompass poor sleep quality, daytime sleepiness, insomnia symptoms, obstructive sleep apnea symptoms and restless leg syndrome symptoms based on self-reported questionnaires on sleep quality and behaviours. More details on sleep disturbance classification are provided in Table S2 Appendix.

### Statistical Analysis

Demographic characteristics were summarized using mean and standard deviation or frequency and percentage, where appropriate. To build the life-course model, the prevalence of midlife and later life risk factors was calculated for the age group 45-64 and 65-85, respectively. Early life risk factors were calculated for all age groups. The prevalence estimates were weighted with inflation weights to account for differences in selection probabilities.^24^

As described elsewhere,^9^ PAF of each risk factor was calculated using Levin’s formula and risk factor overlap was adjusted by applying Norton’s formula^20^, which involves weighting by communalities from principal component analysis (see Table S3 in Appendix for detailed description). Relative risk was taken from the Lancet report^5^ for all risk factors except sleep disturbance, which was derived from a recent meta-analysis of 18 longitudinal studies with an average 9.5 years of follow up assessing the association between sleep disturbance and dementia.^23^

To compare our results with global estimates^5^ and other countries,^7-11^ we obtained the prevalence and PAF from other studies that utilized the Lancet^5^ approach. The life-course model obtained from CLSA was qualitatively compared to the global estimate^5^ and other countries – USA,^7^ India,^9^ Latin America,^9^ China,^9^ New Zealand,^8^ Brazil,^11^ Australia,^10^ and Denmark.^12^ A total of 9 risk factors were available across all the studied countries – less education, hearing loss, hypertension, obesity, smoking, depression, social isolation, physical inactivity, and diabetes – and were measured using similar definitions across studies.

To further explore differences in risk factor distribution by age groups and sex, analyses were stratified by 4 age groups (45-54, 55-64, 65-74, and 75-85) and sex and chi-square tests were conducted. A sensitivity analysis compared changes in prevalence and PAF with different risk factor definitions. Depression was re-operationalized as a 10-item Center for Epidemiologic Studies Depression Scale score of ≥10 to reflect depressive symptom. Excessive alcohol use was re-defined using Canadian guidance, which is drinking ≥7 standard drinks per week.^25^ A five-point Steptoe social isolation index was derived to incorporate cohabitation, social contact and participation into social isolation with a cut-off of ≥3.

## RESULTS

A total of 30,097 participants were included (Table 1). The mean age was 59.7 years (SD 10.3) and 52% were female. The majority were white (94%), living in urban areas (90%), and 74% were married.

**Table 1.** Participant characteristics (Weighted N = 3,812,085)

|  | Overall<br>(n=30,097) |
| --- | --- |
| Age, years | 59.74 (10.30) |
| Age groups |  |
| 45 - 54 | 7,595 (39%) |
| 55 - 64 | 9,856 (31%) |
| 65 - 74 | 7,362 (18%) |
| 75+ | 5,284 (12%) |
| Sex |  |
| Women | 15,320 (52%) |
| Men | 14,777 (48%) |
| Ethnicity |  |
| Non-white | 1,326 (6.2%) |
| White | 28,771 (94%) |
| Education |  |
| < secondary school education | 1,643 (17%) |
| Secondary school graduation | 2,839 (12%) |
| Some post-secondary education | 2,238 (9.1%) |
| Post-secondary degree/diploma | 23,327 (62%) |
| Marital |  |
| Single (never married/lived with a partner) | 2,654 (8.7%) |
| Married/Common-law relationship | 20,651 (74%) |
| Widowed/Divorced/Separated | 6,784 (17%) |
| Income |  |
| <\$20,000 | 1,566 (6.9%) |
| \$20,000 - \$50,000 | 6,360 (23%) |
| \$50,000 - \$100,000 | 9,907 (33%) |
| \$100,000 - \$150,000 | 5,524 (20%) |
| >\$150,000 | 4,799 (17%) |
| Province |  |
| Alberta | 2,957 (10%) |
| British Columbia | 6,254 (28%) |
| Manitoba | 3,113 (7.2%) |
| Newfoundland and Labrador | 2,214 (2.1%) |
| Nova Scotia | 3,078 (3.8%) |
| Ontario | 6,418 (17%) |
| Quebec | 6,063 (32%) |
| Region |  |
| Pacific | 6,254 (28%) |
| Prairie | 6,070 (18%) |
| Central | 12,481 (48%) |
| Atlantic | 5,292 (5.8%) |
| Rurality |  |
| Rural | 2,424 (5.2%) |
| Urban | 26,461 (90%) |
| Peri-urban | 1,212 (4.3%) |

Data shown are mean (SD) or n (%)

### Prevalence

The most prevalent risk factor was later life physical inactivity (83%) (Table 2), followed by later life sleep disturbance (40%), midlife obesity (31%), midlife hypertension (30%), and midlife hearing loss (21%). The least common risk factors were midlife smoking (6.2%) and social isolation (1.6%) in later life. The prevalence of physical inactivity was much higher than in other countries, which ranged from 15.3% in India to 82% in Australia, as shown in Table 3. Smoking and social isolation were noticeably less prevalent than in other countries.

**Table 2.** PAF for 12 modifiable risk factors for dementia in Canada.

|  | Risk factors | RR | Communality<br>(%) | Prevalence<br>(%) | Unweighted<br>PAF (%) | Weighted PAF (%) |
| --- | --- | --- | --- | --- | --- | --- |
| Early life | Less education | 1.6 (1.3, 2.0) | 56.3 | 14.0 | 7.8 | 3.2 (1.9, 4.3) |
| Midlife | Hearing loss | 1.9 (1.4, 2.7) | 52.3 | 21.0 | 15.9 | 6.5 (3.7, 9.3) |
|  | Traumatic brain injury | 1.8 (1.5, 2.2) | 18.3 | 15.0 | 10.7 | 4.4 (3.3, 5.4) |
|  | Hypertension | 1.6 (1.2, 2.2) | 55.4 | 30.0 | 15.3 | 6.2 (2.7, 9.3) |
|  | Excessive alcohol | 1.2 (1.1, 1.3) | 43.5 | 11.0 | 2.2 | 0.9 (0.5, 1.1) |
|  | Obesity | 1.6 (1.3, 1.9) | 57.1 | 31.0 | 15.7 | 6.4 (4.1, 7.7) |
| Later life | Smoking | 1.6 (1.2, 2.2) | 51.7 | 6.2 | 3.6 | 1.5 (0.6, 2.4) |
|  | Depression | 1.9 (1.6, 2.3) | 22.3 | 12.0 | 9.8 | 4.0 (3.2, 4.8) |
|  | Social isolation | 1.6 (1.3, 1.9) | 29.8 | 1.6 | 1.0 | 0.4 (0.2, 0.5) |
|  | Physical inactivity | 1.4 (1.2, 1.7) | 54.1 | 83.0 | 24.9 | 10.2 (6.8, 13) |
|  | Type 2 diabetes | 1.5 (1.3, 1.8) | 40.0 | 13.0 | 6.1 | 2.5 (2.4, 3.3) |
|  | Sleep disturbance | 1.2 (1.1, 1.3) | 24.6 | 40.0 | 7.4 | 3.0 (1.8, 3.8) |
| Combined PAF |  |  |  |  |  | 49.2 (31.1, 64.9) |

Abbreviations: PAF, population attributable fraction; RR, risk ratio

**Table 3.**
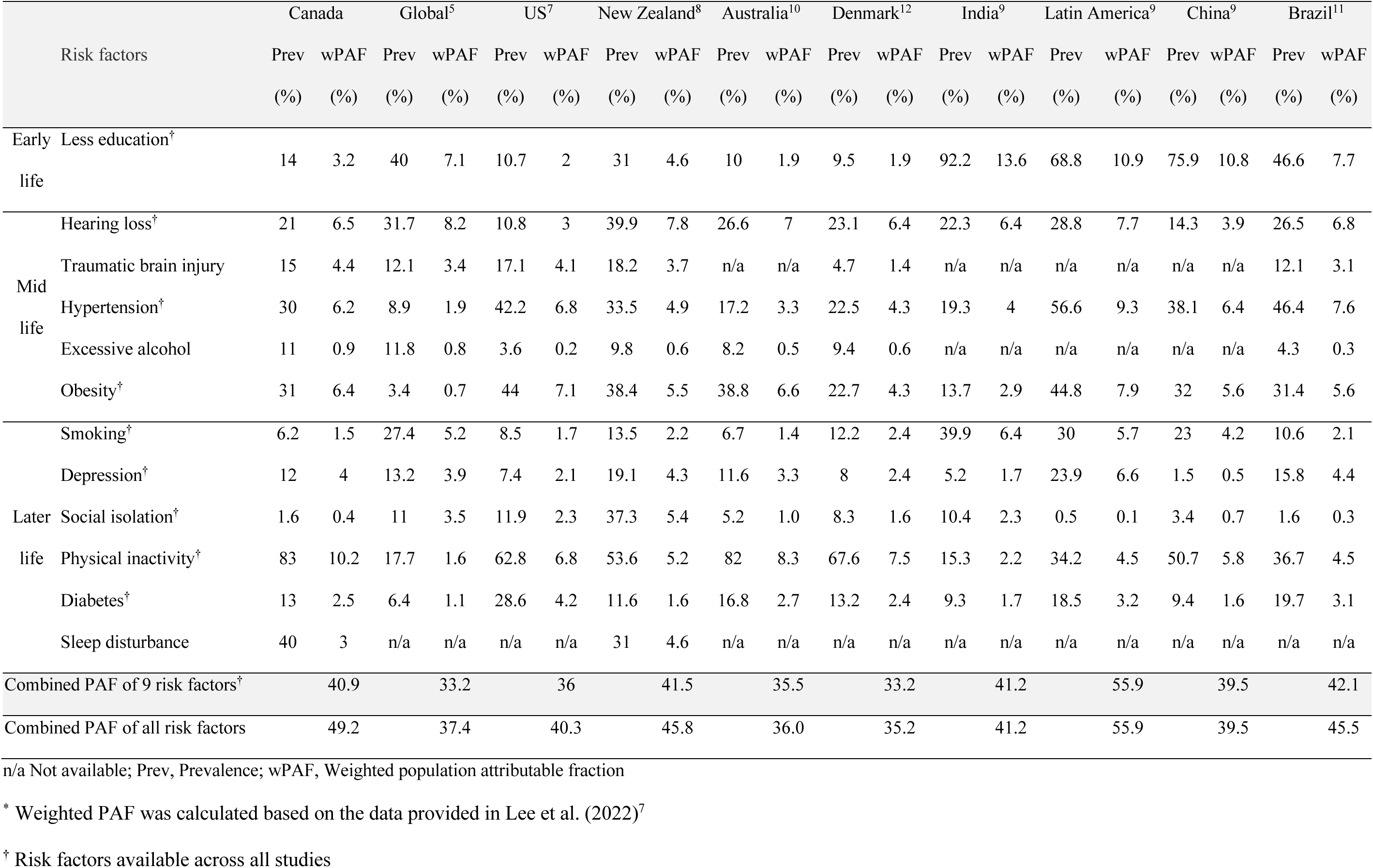
Prevalence and weighted PAF of 12 potentially modifiable dementia risk factors over the life course in Canada compared to the estimates reported in other countries.

### PAF

The weighted individual and combined PAF of all risk factors in Canada are presented in Table 2 and Figure 1. Almost 50% of dementia cases in Canada were attributed to 12 risk factors (PAF 49.2%, 95% CI 31.1% to 64.9%) – nearly 12 points more than the worldwide estimate of 37.4% (Table 3 and Figure S2 in Appendix). The PAF of the nine risk factors available in other studied countries ranged between 33.2% and 55.9%. When using these nine risk factors, the combined PAF in Canada (41%) was higher than in USA (36%), Denmark (33.2%) and Australia (35.5%) but lower than in Latin America (55.9%). The combined PAF of the nine risk factors in Canada was similar to New Zealand, India, China, and Brazil, with a range from 39.5% and 42.1% (Table 3).

**Figure 1.**
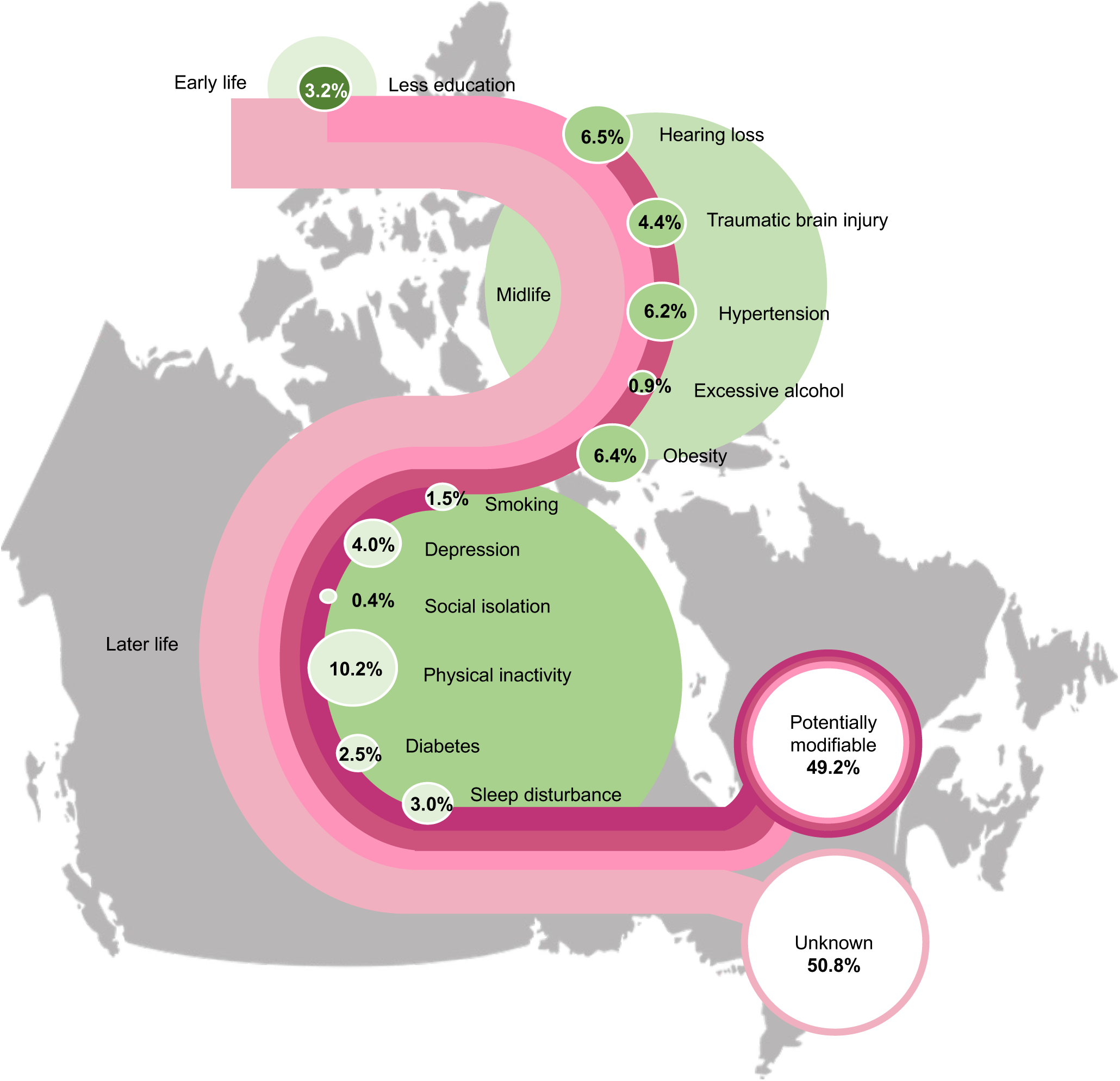
Weighted population attributable fraction for 12 potentially modifiable risk factors for dementia in Canada.

Of the 12 risk factors, later life physical inactivity had the largest weighted PAF in Canada, contributing 10.2% (95% CI 6.8% to 13.0%). The PAF of midlife hearing loss, obesity, and hypertension in Canada were 6.5%, 6.4%, and 6.2% respectively (95% CI 3.7% to 9.2% for hearing loss, 95% CI 4.1% to 7.7% for obesity, and 95% CI 2.7% to 9.3% for hypertension). Later life smoking, midlife excessive alcohol use, and later life social isolation were associated with less than 2% of PAF in Canada. Compared to other countries, Canadian weighted PAFs were smaller for later life smoking and social isolation, while larger for later life physical inactivity, midlife traumatic brain injury and excessive alcohol use (Table 3 and Figure S2 in Appendix). The weighted PAFs for early life less education, midlife hypertension, and later life depression in Canada were larger than in other high-income countries, whereas the weighted PAF for early life less education and midlife hypertension was smaller than low- and middle-income countries. Moreover, compared to low- and middle-income countries, Canada had a larger weighted PAF for midlife obesity.

### By Sex and Age groups

Prevalence of the most common risk factors differed across sexes (Table 4 and Figure S4 in Appendix). Among women, 80.0% had physical inactivity and 20.8% had depression, as compared to 72.8% and 11.8% in men. The prevalence of less education and sleep disturbance was similar between sexes (p>0.05). The prevalence of hearing loss, traumatic brain injury, hypertension, excessive alcohol use, diabetes, and social isolation was higher in men than women (p<0.001).

**Table 4.**
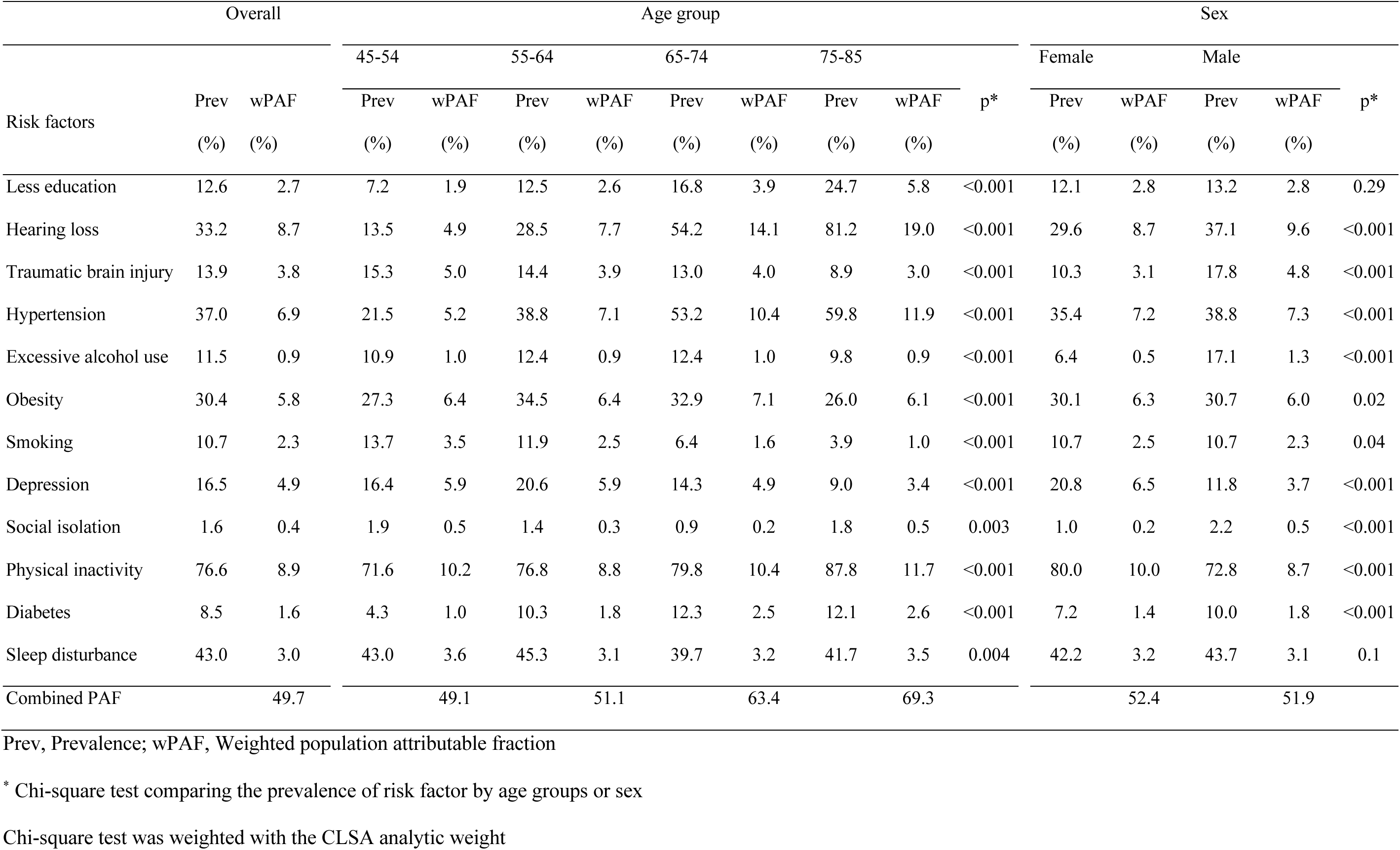
Weighted prevalence and PAF of 12 potentially modifiable risk factors by 4 age groups and sex in Canada.

Overall, the 12 risk factors accounted for 52.4% (95% CI 32.5% to 68.7%) of dementia among women and 51.9% (95% CI 32.2% to 68.0%) among men (Table 4 and Figure S5 in Appendix). The weighted PAF of hearing loss, traumatic brain injury, and excessive alcohol use was higher in men, whereas the weighted PAF of depression and physical inactivity was higher in women.

Prevalence and weighted PAF also varied across age groups (Table 4 and Figure S6 in Appendix). From age 45-54 to 75-85 years, prevalence increased from 13.5% to 81.2% for hearing loss, 7.2% to 24.7% for less education, 21.5% to 59.8% for hypertension, 71.6% to 87.8% for physical inactivity, and 4.3% to 12.1% for diabetes. The highest prevalence of traumatic brain injury (15.3%) and smoking (13.7%) was observed at age 45-54 years, which declined after age 55 years and dropped to 8.9% and 3.9% at age 75-85 years, respectively. Depression prevalence increased from 16.4% in age 45-54 to 20.6% in 55-64 years, then decreased to 9.0% in 75-85 years.

The combined PAF of all 12 risk factors increased with age (Figure S5 in Appendix). The combined PAF were 49.1% (95% CI 30.5% to 65.5%) in age 45-54 years, 51.1% (95% CI 31.4% to 67.6%) in 55-64 years, 63.4% (95% CI 40.7% to 79.4%) in 65-74 years, and 68.2% (95% CI 46.2% to 83.9%) in 75-85 years. Physical inactivity had the highest PAF among ages 45-54 years (10.2%, 95% CI 6.6% to 13.2%) and 55-64 years (8.8%, 95% CI 5.7% to 11.4%), while hearing loss had the largest PAF among ages 65-74 years (14.1%, 95% CI 9.0% to 17.3%) and 75-85 years (19.0%, 95% CI 13.4% to 21.6%). The estimated communality and unadjusted PAFs by age groups and sex are shown in Table S3-S4 in Appendix.

### Sensitivity Analyses

Operationalizing risk factors with different definitions increased the prevalence and PAF, and also changed the rank of risk factors with the largest PAF (Table S5-S7 in Appendix). The prevalence increased to 16% for depression, 29% for excessive alcohol use, and 42% for social isolation. The largest change was observed with social isolation, and its prevalence increased from 1.6% to 42%. The combined PAF increased to 53.2% (95% CI 33.9% to 68.9%). Later life social isolation became the risk factor with the second largest PAF. In older age groups, social isolation had larger PAF than obesity.

## DISCUSSION

Our results suggest that up to 49% of dementia cases in Canada could be delayed or prevented by modifying 12 risk factors including sleep disturbance. To our knowledge, this is the first study estimating the potential population impact of 12 modifiable risk factors for dementia in this country and the first to include sleep disturbance in a life-course model with participants as young as 45 years. The predominant contributors were later life physical inactivity, midlife hearing loss, midlife obesity, and midlife hypertension. In contrast, later life smoking, midlife excessive alcohol use, and later life social isolation had substantially less contribution. Finally, we observed a similar prevention potential in men than women.

Over 80% of Canadians were not meeting physical activity guidelines, and nearly 40% of Canadians had impaired sleeping. Furthermore, 1 in 3 Canadians were obese or had hypertension, and 1 in 5 Canadians showed hearing loss. The estimated prevalence of obesity, physical inactivity, diabetes, and sleep disturbance was similar to previous estimates but our results showed a higher prevalence of less education and hypertension, and a lower prevalence of smoking and hearing loss.^26,27^

Canada’s national dementia strategy includes the promotion of healthy lifestyle to reduce dementia risk, such as initiatives and programs to prevent and manage cardiovascular diseases and promote physical activity.^26,28^ This high prevalence of physical inactivity is found despite sustained national efforts to promote physical activity. It highlights the need to advocate and implement moderate-to-vigorous exercise programs for older adults to help them meet physical activity targets. For instance, aerobic and resistance exercises have been shown to improve cognition in older adults and can be delivered in exercise programs.^29,30^ More innovative approaches which provide paid-for programs in high-trafficked community institutions such as the YMCAs or online training done in the home environment are worth exploration.

There has been extensive development of a national strategic plan for hypertension prevention and control in Canada since 1990s.^31^ However, the prevalence of vascular risk factors, hypertension and obesity, we observed were still relatively higher than other risk factors. While a low prevalence of diabetes may reflect the national strategic plans, such as sodium reduction strategy^31^, it could be also due to not considering blood glucose levels in its definition. The high prevalence of hypertension and obesity underscores the opportunity to improve both vascular and cognitive health.

Despite an unclear underlying mechanism between hearing loss and dementia, a meta-analysis of 8 longitudinal studies found a 19% lower risk of cognitive decline among individuals with corrected versus uncorrected hearing loss (HR 0.81, 95% CI 0.76-0.87).^32^ These findings suggest that dementia prevention programs and primary health care providers should consider encouraging hearing tests and hearing aid use. Compared to other lifestyle strategies, addressing hearing loss faces challenges related to stigma and affordability of hearing aids.^33^ Given its high PAF and associated barriers, there is a pressing need to enhance hearing health care besides promoting patient education.

Sleep disturbance had a substantial contribution to dementia and the high prevalence we found is noteworthy because it can be translated to large potential population health benefits were it to be successfully addressed. Non-pharmacological interventions for sleep disturbance or disorder include sleep hygiene education, physical activity, and in some cases, bright light therapy,^34^ however their effectiveness in preventing cognitive impairment requires further study. Periodically screening sleep impairment in practice will help identify and intervene undiagnosed sleep impairment.

Our age-specific analysis indicated that up to 49% of dementia cases could be prevented if we can modify all 12 risk factors as early as at age 45, and the potential for dementia prevention increased to 69% at age 75. Interestingly, the prevalence of physical inactivity, obesity, hypertension, and sleep disturbance was already high at age 45-54 and all the predominant risk factors identified in the life-course model, except physical inactivity, were from midlife. These results support the recommendation of commencing risk reduction interventions at midlife or earlier, before developing cognitive impairment.^35^.

Our age-specific analysis results align with a study from Chile, which calculated the PAF of dementia risk factors, stratified by sex and 2 age groups (45-64 and ≥65). ^36^ Similarly, we found that physical inactivity, less education, and risk factors related to chronic diseases, such as hearing loss, hypertension, and diabetes, gradually increase with age. The observed decreasing trend for traumatic brain injury, smoking, and excessive alcohol use may imply a decline in risk-taking with aging.^37^

As with other studies ascertaining sex-specific estimates,^36,38,39^ the prevalence of traumatic brain injury, hearing loss and excessive alcohol use were higher in men while depression and physical inactivity were more prevalent in women. Contrary to our results, sex differences in smoking were observed in previous studies.^36,38,39^ Although we obtained a similar potential for dementia prevention in men and women, the observed difference in risk factor profile is important to inform public policy to focus on addressing early risk behaviours, particularly in men.

Importantly, the difference in the risk factor profile we observed by age groups and sex suggests the importance of tailoring dementia prevention programs and strategies within Canada. For instance, dementia prevention programs for middle-aged adults should include education on risk-taking behaviours, while the focus could shift to better management of hearing loss, hypertension, and diabetes in older adults. The programs for older adults should prioritize delivering group exercise or recreational programs. The dementia prevention efforts could be further enhanced by focusing on addressing depression in women and hearing loss in men.

The combined PAF of the nine risk factors that are available across all studies was 33.2% globally and ranged between 33.2% and 55.9% among studied countries. The proportion of potentially preventable dementia cases attributable to these nine risk factors in Canada was similar to studies from low- and middle-income countries, but higher than those from other high-income countries. Latin America reported the highest PAF, using data from 6 Latin American countries.^9^

The risk factor with the highest PAF was less education in middle- and low-income countries, while it was physical inactivity and obesity in high-income countries, suggesting a sociocultural gradient in dementia risk factors. Interestingly, risk factor profiles varied even among high-income countries, highlighting the uniqueness of the Canadian population which could inform policymakers in Canada. For instance, despite cultural similarities between USA and Canada, the prevalence of hearing loss, depression, and alcohol use was 2 to 4 times higher in Canada while USA had 1.5 times or more higher prevalence of obesity, diabetes, hypertension, and social isolation.^7^ Despite geographical distance, the risk factor profile of Canada was fairly similar to Australia and Denmark. This underpins the importance of identifying population-specific risk factor profiles to inform researchers and policymakers on developing dementia risk reduction strategies. International differences in risk factor prevalence could also reflect healthcare and policy contexts. For instance, the relatively lower prevalence of less education and smoking in high-income countries might be explained by the implementation of governmental compulsory education^40^ and tobacco control policies.^41^

The primary strength of our study is the use of CLSA, the largest and well-characterized cohort study in Canada. Its large sample allowed us to explore how risk factor distributions differ from midlife to later life. Our study is the first to show dementia risk factor distributions across 4 age groups beginning at age 45.

There are several limitations. First, some risk factors were defined based on self-reported data and misclassification of risk factors may impact the estimated PAF. For example, depression and sleep disturbance were classified based on self-reported measures, although using valid instruments. Physical inactivity excluded light activities. However, because our sensitivity analyses produced higher estimates of PAF than our main analysis, our estimates probably underestimate PAF. Second, our sample was composed by largely highly educated White Canadians, living in urban settings, which limits generalizability. Third, Levin’s formula assumes no confounding and it produces a biased estimate of PAF when adjusted relative risk is used. Considering that the adjusted relative risk for dementia risk factors tends to be less than the unadjusted relative risk, our obtained PAF may be underestimated.^42^ For age and sex-specific PAF calculation, the relative risk used was from life course-specific measures and may not accurately represent other age groups or sexes. We also acknowledge that the interpretation of the PAF is based on assumptions of causality and the complete eradication of risk factors, which is unlikely to be achieved in real life. However, it can still inform public health intervention strategies on which risk factors to be prioritized. Given the multifactorial nature of dementia, our approach of adjusting for risk factor inter-relationship to combine PAF may not fully account for confounding, interactions among risk factors and its multifactorial nature. Although diet is recognized as an important dementia risk factor^43^ it was not included due to the complexities of analyzing diet. Lastly, while most of the risk factors had similar definitions, caution is needed in comparing the prevalence and PAF across studies.

Our results have critical implications for healthcare professionals, increasing their awareness of dementia risk factors and promoting the screening of prevalent risk factors to facilitate patient’s risk reduction strategies from midlife. Furthermore, the findings could help researchers and policymakers design optimal prevention programs to ease the projected burden of dementia. Although today the *β*-amyloid-targeting therapies in early symptomatic Alzheimer’s disease started to show promising results,^44,45^ there is emerging evidence that the beneficial effect of lifestyle intervention are independent of *β*-amyloid pathology.^46^ The FINGER trial showed that a multidomain intervention including exercise, nutritional counselling, cognitive training, social stimulation, and cardiometabolic management, improved cognition and reduced cognitive decline risk in 1,260 community dwellers at risk of dementia.^47^ The FINGER model is being adapted and tested globally within the World-Wide FINGERS network of multidomain trials for dementia risk reduction^14^, with Canada being at the forefront of such efforts with the CAN-THUMBS UP project.^16^ Findings from the present study can inform the refinement and development of the CAN-THUMBS UP multidomain interventions.

Additionally, combining lifestyle intervention with disease-modifying therapy will offer an even better opportunity to effectively and precisely manage and prevent dementia based on patient’s risk profiles, as demonstrated in diabetes management,^48^ and multidomain lifestyle-based interventions combined with repurposed drugs (metformin) are now being tested in Europe.^49^ Importantly, not all older adults at risk of dementia will be candidates for emerging disease-modifying therapy due to the therapies contraindications and limited generalizability of the current trial results, and therefore lifestyle interventions may be more applicable.^50^ Analyses of the most prevalent combinations of risk factors and risk factor clustering would allow the development of impactful personalized prevention programs.

In conclusion, we determined that nearly up to 50% of dementia cases in Canada could be prevented by modifying 12 risk factors across the life span. The most prominent modifiable risk factors were later life physical inactivity, midlife obesity, midlife hypertension, and midlife hearing loss, whereas midlife excessive alcohol use, later life smoking, and later life social isolation had substantially less contribution.

## Data Availability

Data are available from the Canadian Longitudinal Study on Aging (www.clsa-elcv.ca) for researchers who meet the criteria for access to de-identified CLSA data.

## ACKNOWLEDGEMENTS

Surim Son was awarded Alzheimer Society London and Middlesex Doctoral scholarship, Ontario Scholarship Award, and CCNA Trainee Synapse Challenge Funding.

## Authors contribution

Son had full access to all the data in the study and took responsibility for the integrity of the data and the accuracy of the data analysis.

Concept and design: Son, Speechley, Montero-Odasso

Acquisition, analysis, or interpretation of data: Son, Speechley, Zou, Montero-Odasso

Drafting of the manuscript: Son

Revised the manuscript for important intellectual content: All authors

Statistical analysis: Son

Administrative, technical, or material support: Son, Speechley, Montero-Odasso

Supervision: Speechley, Zou, Montero-Odasso

## Statement of Informed Consent

All participants of CLSA study provided informed consent.

